# An effective COVID-19 response in South America: the Uruguayan Conundrum

**DOI:** 10.1101/2020.07.24.20161802

**Authors:** Pilar Moreno, Gonzalo Moratorio, Gregorio Iraola, Álvaro Fajardo, Fabián Aldunate, Marianoel Pereira-Gómez, Paula Perbolianachis, Alicia Costábile, Fernando López-Tort, Diego Simón, Cecilia Salazar, Ignacio Ferrés, Florencia Díaz-Viraqué, Andrés Abin, Mariana Bresque, Matías Fabregat, Matías Maidana, Bernardina Rivera, María E. Cruces, Jorge Rodríguez-Duarte, Paola Scavone, Miguel Alegretti, Adriana Nabón, Gustavo Gagliano, Raquel Rosa, Eduardo Henderson, Estela Bidegain, Leticia Zarantonelli, Vanesa Piattoni, Gonzalo Greif, María E. Francia, Carlos Robello, Rosario Durán, Gustavo Brito, Victoria Bonnecarrere, Miguel Sierra, Rodney Colina, Mónica Marin, Juan Cristina, Ricardo Ehrlich, Fernando Paganini, Henry Cohen, Rafael Radi, Luis Barbeito, José L. Badano, Otto Pritsch, Cecilia Fernández, Rodrigo Arim, Carlos Batthyány, Interinstitutional COVID-19 Working Group.

**Author notes:** These authors contributed equally to this work (Pilar Moreno, Gonzalo Moratorio, Gregorio Iraola).

## Abstract

**Background:** South America has become the new epicenter of the COVID-19 pandemic with more than 1.1M reported cases and >50,000 deaths (June 2020). Conversely, Uruguay stands out as an outlier managing this health crisis with remarkable success.

**Methods:** We developed a molecular diagnostic test to detect SARS-CoV-2. This methodology was transferred to research institutes, public hospitals and academic laboratories all around the country, creating a “*COVID-19 diagnostic lab network*”. Uruguay also implemented active epidemiological surveillance following the “Test, Trace and Isolate” (TETRIS) strategy coupled to real-time genomic epidemiology.

**Results:** Three months after the first cases were detected, the number of positive individuals reached 826 (23 deaths, 112 active cases and 691 recovered). The Uruguayan strategy was based in a close synergy established between the national health authorities and the scientific community. In turn, academia rapidly responded to develop national RT-qPCR tests. Consequently, Uruguay was able to perform ∼1,000 molecular tests per day in a matter of weeks. The “*COVID-19 diagnostic lab network*” performed more than 54% of the molecular tests in the country. This, together with real-time genomics, were instrumental to implement the TETRIS strategy, helping to contain domestic transmission of the main outbreaks registered so far.

**Conclusions:** Uruguay has successfully navigated the first trimester of the COVID-19 health crisis in South America. A rapid response by the scientific community to increase testing capacity, together with national health authorities seeking out the support from the academia were fundamental to successfully contain, until now, the COVID-19 outbreak in the country.

## INTRODUCTION

By mid-2020, Latin America became the epicenter of the COVID-19 pandemic, with more than 1.1M reported cases and 50,000 deaths. In this context, Uruguay stands out as an outlier surpassing the situation with remarkable success. Uruguay is a 3.5- million-people country located in the Atlantic coast of South America. Two rivers (Río de La Plata and Rio Uruguay) establish its border with Argentina to the west, and a long, dry border of almost 1000 km conforms its northeastern limit with Brazil, currently the second country with the highest prevalence of COVID-19 in the world (Worldometers.info/coronavirus). More than 50% of the population lives in Montevideo, Uruguay’s capital city, and its metropolitan area. The other half of the population is distributed among several cities, generally not exceeding 100,000 inhabitants, and the countryside. Additional unique factors compared to other countries, in particular with Argentina and Brazil, are, for example, that there is only one major international airport, located at the periphery of Montevideo, and one major international harbor also located within the capital.

To understand the management of COVID-19 in Uruguay it is important to highlight aspects of its health system. Uruguay has developed a universal access health integrated system with public and private medical centers offering widespread access to all the medical services and procedures, including intensive care. Before SARS-CoV-2 was first detected in Uruguay (March 13^th^ 2020), the health system had 19 intensive care beds per 100,000 inhabitants. This figure positioned the country in between countries with the highest number of ICU beds (approximately 30 beds per 100,000 like Germany and Austria), and countries such as France, Spain or Italy with 11, 10, and 9 beds per 100,000 inhabitants, respectively. Of note, Uruguay was able to increase this number significantly reaching 24 intensive care beds per 100,000 inhabitants just two months after the first COVID-19 cases were detected.

At the time of COVID-19 emergence on March 13^th^, Uruguay decided to base its strategy on two main lines of action that we understand underlie the so far successful management of the epidemic. As opposed to other countries, Uruguay had a rapid, seemingly exaggerated, response declaring a health state of emergency upon detection of the first cases. This included closing borders, shutting down schools and any activity that may lead to agglomeration (pubs, commercial centers, churches, entertainment venues), among other actions. The government also exhorted personal responsibility for controlling the virus through voluntary/self-confinement with no mandatory lockdown (a summary of key factors is shown in Fig. 1). In parallel the government sought for advice, support and help from the local scientific system.

**Figure 1.**
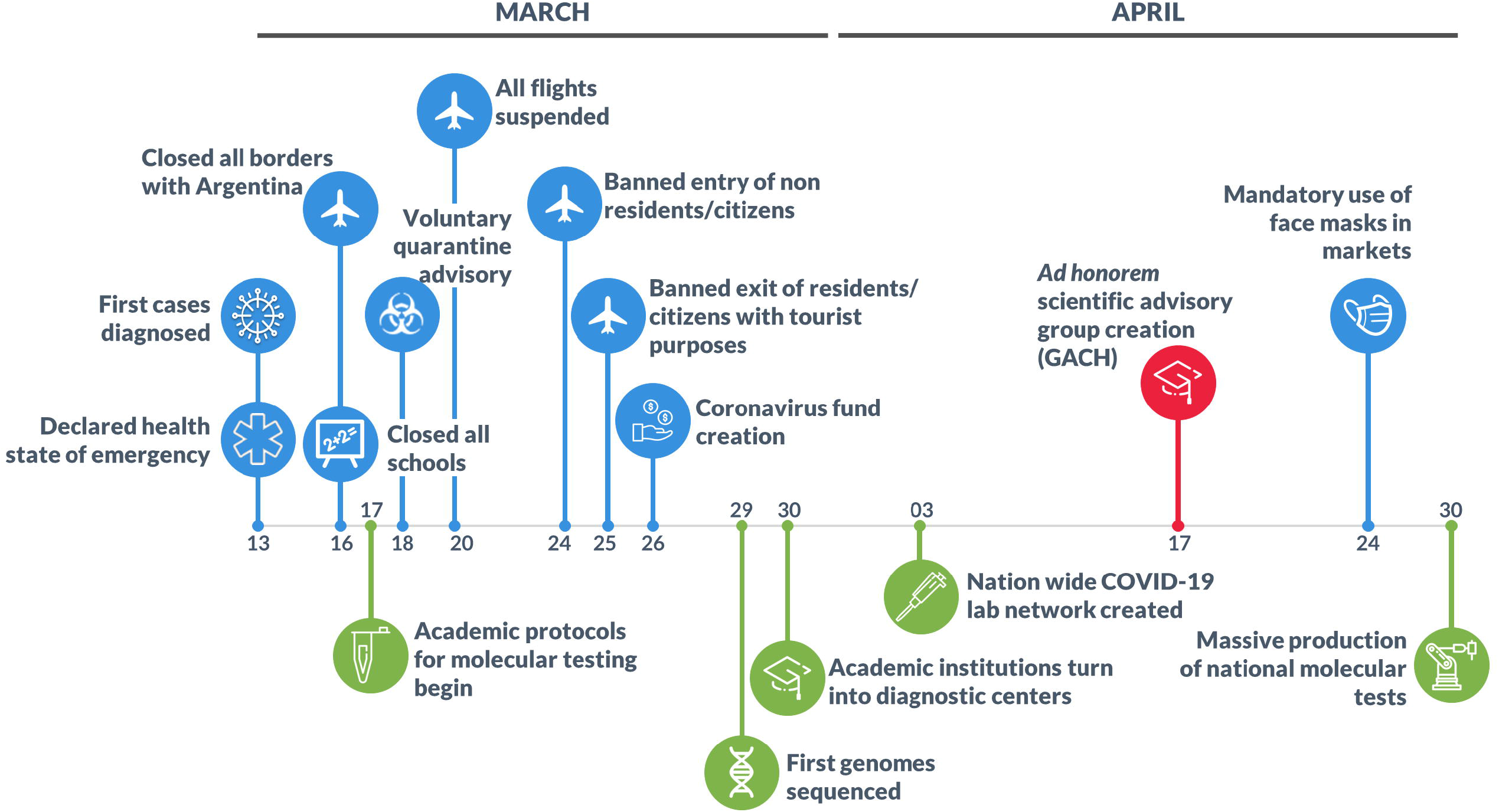
Timeline of main decisions during the first two months of the COVID-19 crisis. Political decisions are shown in blue, Academic/Scientific contributions are shown in green and the *ad honorem* Scientific Advisory Group (GACH) creation is shown in red.

By early March 2020, Uruguay had access to only 200 PCR-based diagnostic assays, a worrisome factor in the face of the incoming pandemic, especially in an international context of imminent shortage of commercial diagnostic kits and reagents as stressed by the World Health Organization (WHO). As a result of a first pivotal meeting and in synergy with the Ministry of Public Health, the Uruguayan scientific and medical ecosystem decided to develop a national response based on four key aspects: i) to locally develop diagnostic resources, ii) to expand the diagnostic capacity by creating a national network of COVID-19 diagnostic labs, iii) to deploy epidemiological approaches and real-time genomic surveillance to track the transmission of the virus and to help contain potential outbreaks. Moreover, a month later an *ad honorem* Scientific Advisory Group (GACH) was created to advice and report directly to the government (see Fig. 1).

Almost three months after the first cases, a period of time in which most countries enter the exponential phase, Uruguay has never entered into such complex phase. In this work, we aim to describe and discuss the key scientific approaches and tools that have been important milestones in the management of the COVID-19 crisis in Uruguay.

## METHODS

### Plasmids and RNA

The *N*, and *Orf 1b* sequences of SARS-CoV-2 were PCR amplified and cloned in TOPO vector, according to instructions provided by the manufacturers (Invitrogen). For *in vitro* transcription, both TOPO-N and TOPO-Orf 1b were linearized with *SpeI* and then transcribed using T7 RNA polymerase (Thermo Fisher). The *in vitro* transcribed RNA was DNase treated, purified with TURBO DNA-free™ Kit (Thermo Fisher) and then was subjected to a tenfold serial dilution until the concentration was between 20 and 15 copies/µl in DEPC-treated water.

### Development of a Molecular SARS-CoV-2 Diagnostic kit: *COVID-19 RT-PCR Real TM Fast-HEX/Cy5 Kit*

Primers and probes targeting conserved regions in the nucleocapsid (N) and the Orf 1b viral genes were adapted and optimized, together with RNase P human gene as a sampling control. These primer/probe sets were subjected to BLAST searching to ensure that they did not align to sequences other than SARS-CoV. The *COVID-19 RT-PCR Real TM Fast-HEX/Cy5* kit is based on the standard hydrolysis probe system (TaqMan® Technology). The SARS-CoV-2 specific probes are labeled with the FAM fluorophore, while the RNAse P probe is custom labeled with either HEX or CY5 fluorophores depending on the equipment requirements. The sampling control is essential to identify possible PCR inhibition, to measure RNA and to confirm the integrity of the PCR run. The primers and probes were synthesized by Integrated DNA Technologies (IDT) and purified by high-pressure liquid chromatography. The OneStep Reaction mix was prepared using 5 µl of RNA samples, 5 µl of 4 x TaqMan Fast Virus 1-Step Master Mix (Thermo Cat. No. 4444436), with 0.25 µM of each N primers, 0.06 µM of each N probe, 0.3 µM of each RNAsaP primer, 0.1 µM of RNAsaP probe and molecular biology grade water to a final volume of 20 µl. The RT-qPCR assays were performed using either Step One Plus, ABI 7500, QuantStudio™ 3, QuantStudio™ 5 (Applied Biosystem) and Rotor Gene 3000/6000 (Corbett Research). The full protocol is available in: http://www.atgen.com.uy/sitio/wp-content/uploads/2020/05/Inserto-71-v2-HEX.pdf. This kit was evaluated using serially diluted *in vitro* transcribed RNA, as described above, from the *N* and *Orf 1b* genes of SARS-CoV-2 as a positive control.

The analytical sensitivity of the test was defined as the lowest concentration of viral *N* gene RNA that could be reliably detected with 95% confidence. This was assessed using synthetic RNAs from de *N* gene with known copy number. Specificity studies were assessed through *in silico* sequence comparison analyses.

### Molecular Diagnosis of SARS-CoV-2

Throat swabs and others respiratory samples were collected for extracting SARS-CoV-2 RNA both from presumptively infected patients and samples obtained from random tests. Viral RNA was extracted using different RNA extraction kits depending on their availability. The following kits were used in the period of time described in this article: “Monarch Total RNA Miniprep Kit” (New England Biolabs); “QIAamp Viral RNA mini kit” (QIAgen) and Viral DNA/RNA Isolation Kit (lifeRiver). All reagents were used according to the manufacturer’s protocol. Also, when RNA extraction kits were unavailable, TRIzol Reagent was used (Invitrogen). The extracted RNA was used then for the qualitative detection of SARS-CoV-2 using de national kit “*COVID-19 RT-PCR Real TM Fast-HEX/Cy5 Kit”* according to the manufacturer’s protocol (ATGen, Institut Pasteur de Montevideo, Universidad de la República). A Ct-value less than 35 was defined as a positive test result, and a Ct-value of 40 or more was defined as a negative test result. A medium load, defined as a Ct-value of more than 35 to less than 40, required confirmation by retesting. This SARS-CoV-2 molecular detection protocol was transferred to research institutes; public hospitals and academic laboratories all around the country.

### Effective reproduction number (Re) calculation

Re was estimated using a discretized generation time distribution assuming a shifted gamma distribution (discr_si function) implemented on EpiEstim package^1–3^, with serial interval (k) ranging from 0 to 30, mean µ = 3.95 and standard deviation σ = 4.75 as parameters^4,5^. Data handling and plots were performed in R^6^; dates were managed with the Lubridate package^7^ and violin plots using the Vioplot package^8^.

### Genomic analysis

A global collection of complete and high-quality SARS-CoV-2 genome sequences were retrieved from the GISAID (http://www.gisaid.org) database on 2^nd^ of June. A list of sequences and acknowledgments to the originating and submitting labs is presented in Supplementary Table 1. We analyzed this dataset using the Augur Toolkit version 6.4.2^9^. Briefly, genome sequences were aligned using MAFFT^10^ and phylogeny was reconstructed with IQ-TREE^11^. Clades were defined according to the current classification of Nextstrain (www.nextrstrain.org). SARS-CoV-2 genomes from cases reported in Uruguay were sequenced with different technologies, including Illumina and the ARTIC Network protocol using the MinION platform (Oxford Nanopore).

## RESULTS

### Epidemiological status

Three months after the first detected COVID-19 cases the total number of positive cases was 826, with 23 deaths (7 deaths per 1M people), 112 active cases (with only 4 patients requiring intensive care) and 691 recovered individuals. Altogether, 87% of all cases were outpatients, 4% were assisted in intensive care units (ICUs), and the remaining 9% (n = 74) required standard hospitalization. Comorbidities (including cardiovascular disease, obesity, diabetes, chronic pulmonary obstructive disease, kidney diseases, among others) were overrepresented in patients that were admitted to hospitals (71%) compared to ambulatory patients (27%) (Table 1 and 2). Of note, and consistently with observations made worldwide^12^, men appeared to be more affected than women with 5.9% of men requiring critical care assistance vs. 1.7% of women (Table 1).

**Table 1.** Demographic description of confirmed COVID-19 cases and ICU requirement.

| Age - n (%) [ICU] | Female | Male | Total |
| --- | --- | --- | --- |
| <15 | 15 (1.8) [0] | 8 (1) [0] | 23 [0] |
| 15-24 | 51 (6.2) [0] | 37 (4.5) [0] | 88 [0] |
| 25-34 | 88 (10.7) [0] | 104 (12.6) [0] | 192 [0] |
| 35-44 | 62 (7.5) [1] | 63 (7.6) [0] | 125 [1] |
| 45-54 | 51 (6.2) [0] | 59 (7.1) [5] | 110 [5] |
| 55-64 | 70 (8.5) [2] | 68 (8.2) [6] | 138 [8] |
| 65-74 | 35 (4.2) [2] | 37 (4.5) [8] | 72 [10] |
| >75 | 49 (5.9) [2] | 29 (3.5) [5] | 78 [7] |
| **TOTAL** | **421 (51) [7]** | **405 (49) [24]** | **826 [31]** |

**Table 2.**
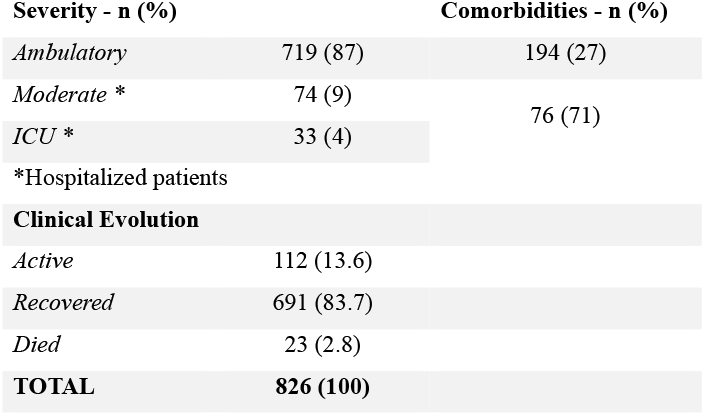
Clinical characteristics of confirmed COVID-19 patients.

The accumulated number of COVID-19 cases in Uruguay never reached an exponential phase. Case number peaks correspond to four isolated outbreaks with less than 40 cases per day each (Fig. 2). The first outbreak occurred at the same time as the first reported cases and was associated with a social event (SE). A second outbreak happened in a mental health care hospital (MH), and a few weeks later a third outbreak occurred in a Residential Care for the Elderly (RCFE) facility. Finally, an outbreak occurred at a city of Rivera (RA), located at the dry border with Brazil, the number two country with the highest number of cases worldwide. Uruguay rapidly contained these outbreaks. Several factors might explain their successful management, and the course of the epidemic in the country, including the rapid tracking and isolation of people that had contacts with confirmed cases. Importantly, that effort was based on the ability of the National Direction of Epidemiology to rapidly confirm positive cases in the first place, thus following the WHO’s recommendation to “test, trace and isolate” (TETRIS). This end could have been, however, complicated by the country’s limited testing capacity at the start of the epidemic coupled with the international shortage of supplies.

**Figure 2.**
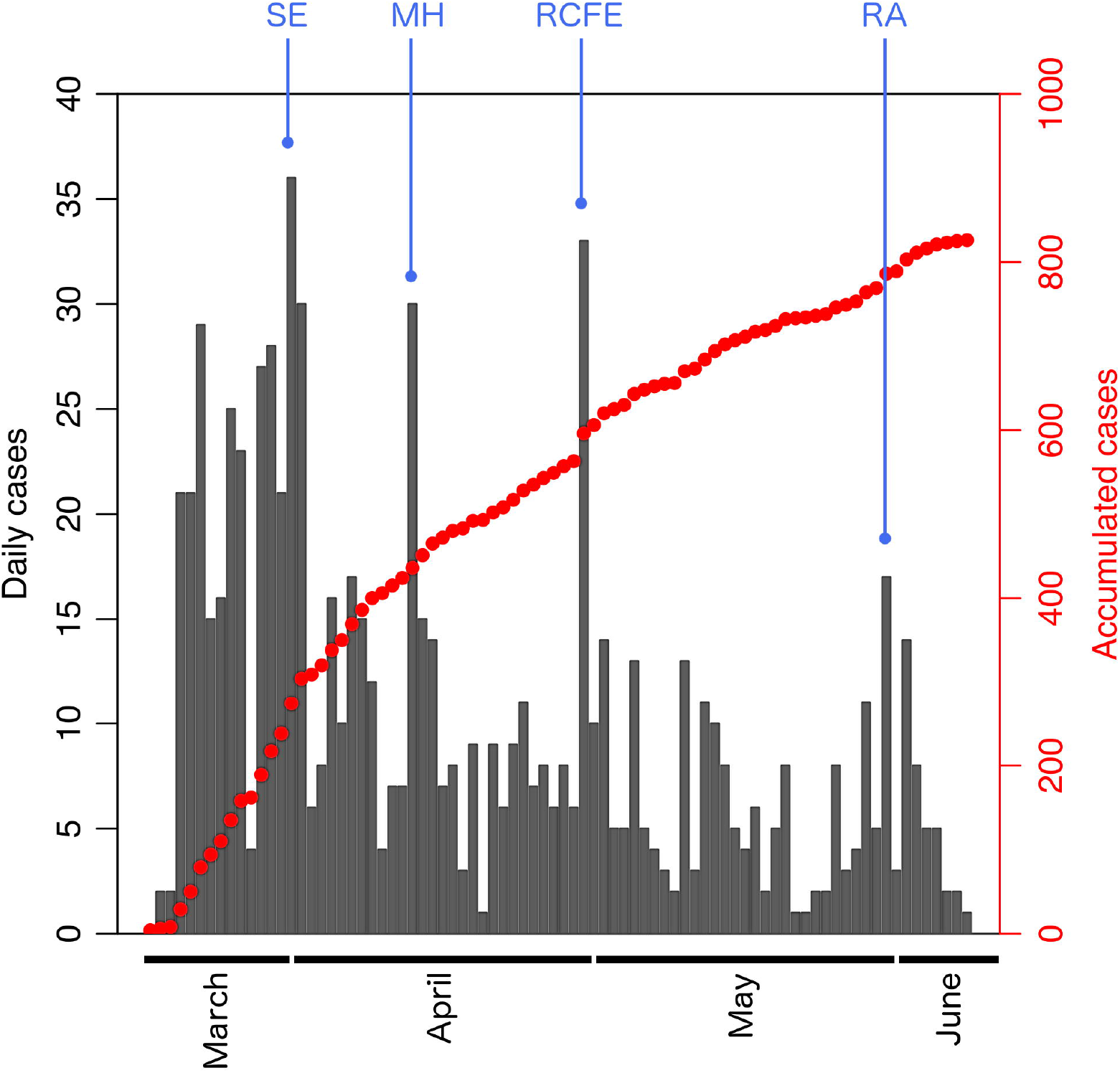
Case evolution since COVID-19 emergence in Uruguay. Red dots show the number of accumulated cases. Black bars point out number of daily cases. Principal outbreaks are shown in blue: Social Event (SE), mental health care hospital (MH), Residential Care for the Elderly (RCFE) facility and Rivera city (RA).

### Development of a national molecular diagnostic test

For the successful implementation of the TETRIS strategy, performing as many molecular tests as possible is a key factor. To achieve this, we first analyzed all molecular protocol available for COVID-19 diagnosis recommended by the WHO. These methods are all based on reverse transcriptase real-time PCR (RT-qPCR) detection of different viral genomic regions^13^. The protocols require several reactions to be carried out per patient, thus increasing the volume and number of different types of reagents required, as well as, the chance of error. With an end goal of becoming external supplier-independent, and to save time and resources, we developed a molecular test based on a multiplex OneStep RT-qPCR assay to detect SARS-CoV-2. To do this, we adapted and optimized a series of primers and probes to target two conserved viral regions, the *nucleocapsid* (*N*) and the *Orf 1b* viral genes, together with the *RNase P* human gene as sampling control (see Methods). The analytical sensitivity of our SARS- CoV2 Kit is 15 copies/μl of viral RNA with a confidence ≥95% (see supplementary Fig.1). This concentration therefore represents the kit’s detection limit. Regarding specificity, the “COVID-19 RT-PCR Real TM Fast-HEX/Cy5” detects all SARS-CoV-2 viral RNA sequences described so far for South America and worldwide.

### Creation of a nation-wide diagnostic network

The second step was to increase our testing capacity by establishing a network of laboratories for SARS-CoV-2 diagnosis. This “*COVID19 diagnostic labs network*” was set up in research institutes, public hospitals and academic laboratories throughout the country. This was only possible because of the remarkably collaborative effort of the entire scientific community, working together in technology transfer and equipment sharing, managing to set up diagnostics laboratories from scratch in a few weeks since the first cases were detected. Importantly, and in line with this effort, the national health authorities enabled research institutions to perform clinical diagnosis for COVID-19, following international standards. As a result, Uruguay was able to dramatically increase its diagnostic capacity from 205 molecular tests per day in March to more than 800 by the end of May (Fig. 2; red line). Notably, the “*COVID-19 diagnostic labs network*” performed more than 54% of the molecular tests in the country, reaching almost 100 tests per confirmed COVID-19 case per day (Worldometers.info/coronavirus). The development of a national molecular diagnostic kit and the creation of a nation-wide diagnostic network represented the fundamental tools that enabled the application of continuous epidemiological surveillance to trace and isolate the positive cases and their contacts, which was essential to decrease the effective reproduction number (Re) (Fig. 3).

**Figure 3:**
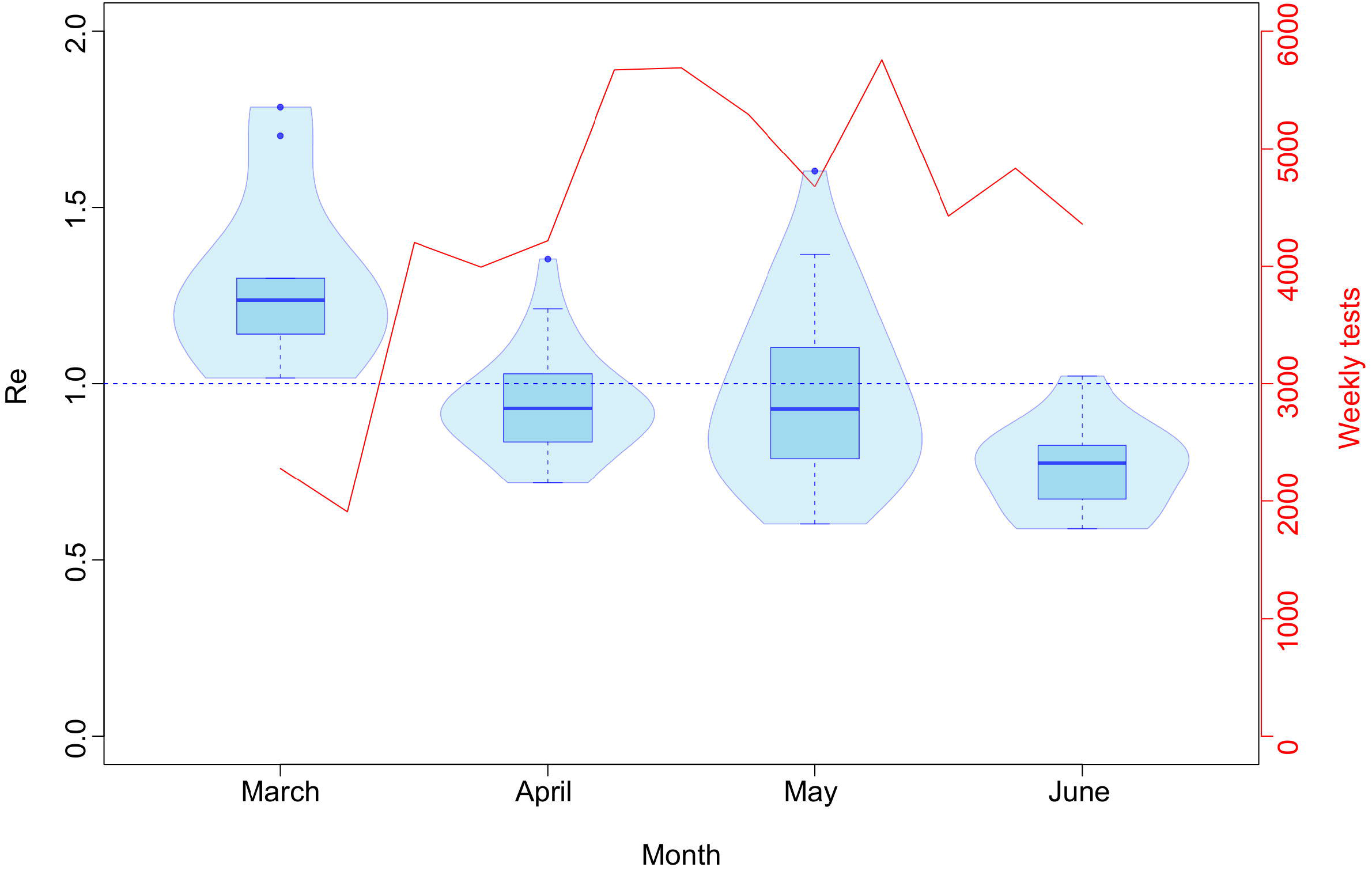
Distribution of the effective reproduction number (Re) values and COVID-19 tests performed by month. The left axis shows the Re value. All values in March are above Re = 1 (dashed blue line), while the majority of values in April-June are below this threshold. Also shown, weekly tests from March 13th to June 13th (secondary axis, in red).

### Rapid contact tracing

To better understand the local transmission dynamics of SARS-CoV-2 and the epidemiological relationship among cases, we analyzed contact tracing data and transmission chains from the four major outbreaks registered in the country (Fig. 4). Interestingly, we noted that outbreak size decreased from March to June, being SE the first and biggest outbreak (n = 190) and RA the last and smallest (n = 39) to date. Also, no transmission chain was longer than 5 generations and most of them (> 50%) were blocked in the second generation. Indeed, longest transmission chains (5 generations) were only registered during the first two outbreaks (SE and MH) and represented ∼ 1% of cases.

**Figure 4.**
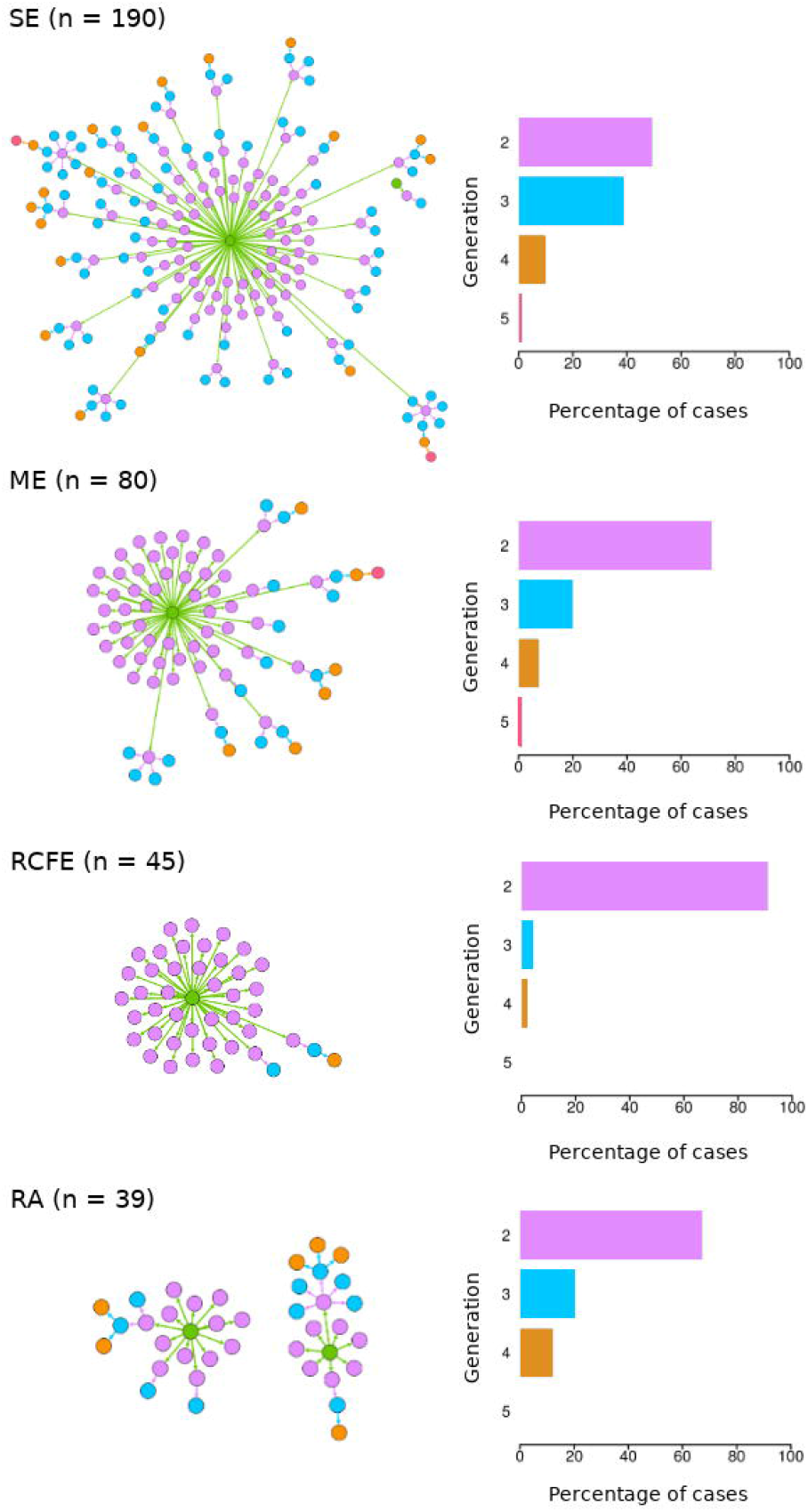
Local transmission chains of four major outbreaks. Each dot represents a confirmed COVID-19 case and edges between dots represents a confirmed epidemiological contact. Each dot is colored according to the transmission generation. Bar plots show the percentage of cases assigned to each transmission generation.

### Real-Time genomic surveillance

At the time of writing this article, Uruguay ranked 5^th^ in the world in the percentage of publicly available SARS-CoV-2 genome sequences relative to the total number of reported COVID-19 cases (considering those countries that have reported at least 500 cases) (Fig. 5A). This was the product of implementing a real-time genomic surveillance approach to characterize genetic variants that arrived in the country and subsequently caused major outbreaks.

**Figure 5.**
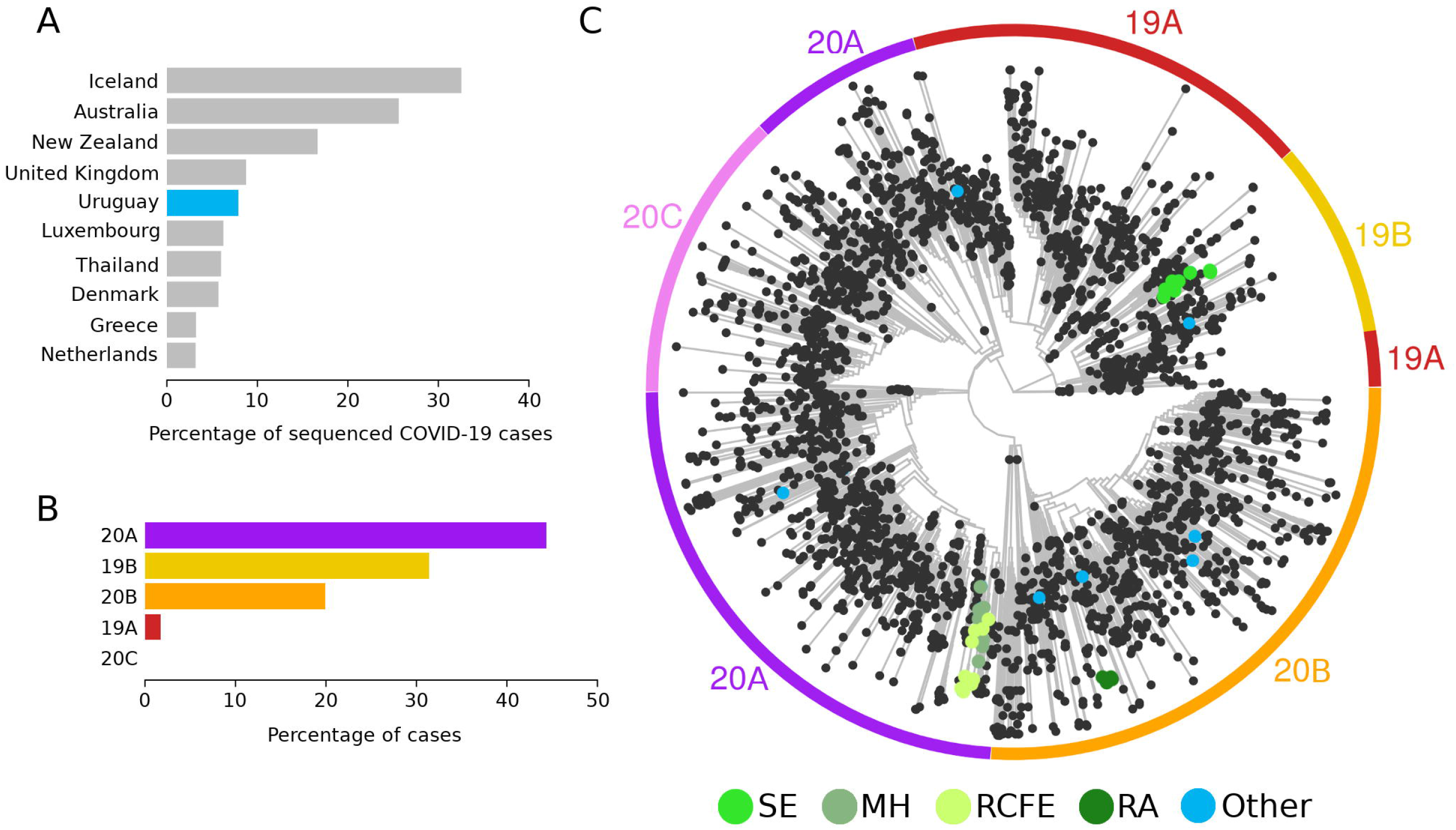
Genomic epidemiology in Uruguay. A) Top 10 countries that have sequenced and publicly released more SARS-CoV-2 genomes relative to the total number of reported COVID-19 cases. B) Distribution of lineages identified in sequenced SARS-CoV-2 genomes from Uruguay. C) Global phylogeny of SARS-CoV-2 highlighting genomes sequenced from SARS-CoV-2 reported in Uruguay.

Our analysis revealed multiple introductions of SARS-CoV-2 into Uruguay and that 4 out of the 5 major SARS-CoV-2 lineages identified worldwide have circulated in the country (Fig. 5B and 5C). Importantly, the application of this genomic epidemiology analysis evidenced that the RCFE outbreak was originated from genetic variants that had already caused the MH outbreak, suggesting a previously unseen epidemiological link between these outbreaks. Remarkably, the genetic variants causing the MH, RCFE and RA represent the recent introduction and local expansion of lineages 20A and 20B (>50% of sequenced cases), which harbor the D614G mutation in the spike protein coding gene. This mutation has been recently proposed to increase viral transmission^14^; hence it would potentially represent an important genetic marker to guide improved disease control strategies.

The application of genomic epidemiology was possible due to the availability of installed second- and third-generation sequencing technologies (like Illumina and Oxford Nanopore) in national academic centers and the creation of an inter-institutional research group focused on the generation and analysis of genomic sequences.

### Increasing testing capacity under viral low-prevalence conditions: matrix pooling

Given the aforementioned strategy, Uruguay managed to keep a low COVID-19 prevalence in the first three month of the outbreak, which allowed us to carefully implement different pooling strategies. Testing samples from many individuals at once allows saving time and reagents to continue increasing our testing capabilities^15^. In the “Matrix pooling” strategy individual samples are arranged in a two-dimensional 10 x 10 grid. Samples are pooled by row and by column and these pools of 10 are tested in the first stage of the algorithm. Individual testing needs to be performed at the sample in the intersection of positive rows and columns. The “Matrix pooling 10×10” strategy allows to analyze 1,000 samples performing only 200 molecular tests. Therefore, this configuration reduces the required number of tests by 80% when compared to individual testing, further increasing our national testing capacity.

## DISCUSSION

The global response to the COVID-19 pandemic has included a wide range of strategies, some even questioning the general guidelines suggested by the WHO. These different decisions range from implementing a mandatory lockdown in some countries, to allowing the spread of the virus in an attempt to achieve herd immunity, to almost disregarding the virus as a true threat to public health. Importantly, these different strategies have largely impacted in the results obtained by each country and worldwide.

Despite being one of the last continents to be hit by the virus and having had the opportunity to observe the outcomes in China, South Korea, and Europe, South America also showed a wide spectrum of responses. At the time of writing this article Uruguay had documented the lowest number of COVID-19 cases per capita in South America, if not in the entire western hemisphere. Uruguay ranked 2^nd^ in South America with the lowest number of deaths per millionth of population (Worldometers.info/coronavirus). This was achieved despite sharing more than 1,000 km of a dry border with Brazil –the country with the world’s second-highest incidence and mortality rates of COVID-19, only behind the United States of America. So far, Uruguay has managed to control the COVID-19 health crisis successfully and is being observed worldwide as a model case (for example^16^).

While the specific demographics of Uruguay could also be considered, our analysis identifies two main aspects that are central and perhaps distinctive. First, the national health authorities sought the support and advice from the scientific system while adopting a rapid shock response (e.g. closing schools and limiting social gathering, albeit avoiding a mandatory quarantine). Second, and aiming to follow the WHO TETRIS guides, the scientific and medical community played an active role in finding national solutions in, at least, three main areas: i) the development of a SARS-CoV-2 RT-qPCR tests, ii) increasing the nation-wide testing capacity and, iii) performing an epidemiological surveillance to track and isolate positive cases while monitoring the circulation of the virus by sequencing technologies.

The spread of the virus worldwide has changed dramatically in the last few months. A frightening fact, for example, is that while it took almost three months to reach the first 500,000 positive cases, 500,000 new cases are now reported in a matter of weeks or even days (Worldometers.info/coronavirus). Thus, controlling the exponential spread of SARS-CoV-2 has been an enormous challenge. In this context, the fact that Uruguay was able to avoid entering an exponential phase of disease contagion in the first three months despite having a number of outbreaks, is a remarkable point. Our analysis shows that one key aspect was the reduction in the effective reproduction number (Re) which is reflected in that most local transmission chains have been controlled at the second generation of contacts. This was due to a massive testing strategy, allowing the rapid detection of COVID-19 cases, both symptomatic and asymptomatic, based on the national development of a RT-qPCR kit and an extensive network of diagnostic laboratories created by the scientific system and supported and encouraged by the national health authorities. In fact, Uruguay was able to reach a number of PCR-based tests per day that compares and sometimes even surpasses top-ranking economies (300 tests per million people per day). Another key factor was the effectiveness of the epidemiological control teams of the Ministry of Public Health who reconstructed the epidemiological changes and followed up most cases on a one to one basis. This in turn allowed to track and quarantine all the people that contacted confirmed positive cases thus limiting the spread of the virus. Importantly, as of to date, we have not reached saturation in our testing and epidemiological surveillance capacity. In addition, the dynamics of the virus transmission was monitored in a real-time fashion through genomic sequencing. These data were also instrumental to understand the different entry events and outbreaks.

Another important aspect that may have facilitated containing the spread of the virus has to do with the characteristics of the national health system. In Uruguay, the first level of medical attention is supported by family physicians that typically assist patients at their home (a strategy that is being strengthened due to the COVID-19 epidemic) therefore reducing the number of potentially infected individuals entering hospitals and clinics. A direct consequence of all these measures has been the fact that we have not saturated neither the epidemiological surveillance system nor the health system, and particularly the ICUs that were specially prepared to deal with the epidemic, even significantly increasing the number of beds available in the system. Importantly, Uruguay needs to keep all the measures and continue exploring innovative strategies to improve the TETRIS. This is especially relevant since mathematical modeling indicates that containment measures can become saturated, giving rise to an epidemic threshold. After that point, the disease begins to grow rapidly in an exponential phase and the previously effective measures become ineffective^17^.

In summary, we firmly believe that the management of the health crisis has been supported by a hitherto unseen interaction between the national health authorities and the scientific academy. This relationship has been instrumental to capitalize on having scientists with different expertise and covering different areas such as virology, epidemiology and molecular biology, many of whom collaborated *ad honorem* to allow the different scientific developments described here. Notably, this interaction has contributed to make decisions based on solid scientific evidence, a paradigm shift that is proving successful so far.

## Data Availability

All data is available upon request

## ACKNOWLEDGMENTS

We thank the Institut Pasteur de Montevideo supporting team for helping us to continue working during the health state of emergency. This work was supported by FOCEM (Fondo de Convergencia Estructural del Mercosur) grant COF 03/11, ANII (Agencia Nacional de Investigación e Innovación), IDB (Inter-American Development Bank), Udelar (Universidad de la República), Institut Pasteur de Montevideo, INIA (Instituto Nacional de Investigación Agropecuaria), IIBCE (Instituto de Investigaciones Biológicas Clemente Estable), BSE (Banco de Seguros del Estado), ATGen and private donations. We deeply appreciate the collaboration and support received by different academic and scientific institutions.

## Supplementary Figure Legends

**Supplementary Figure 1. Analytical sensitivity of the “COVID-19 RT-PCR Real TM Fast-HEX/Cy5” kit**.

A) Amplification plots of sample serial dilutions. Negative samples for SARS-CoV-2 were mixed with known copy numbers of synthetic RNA containing target viral region. 1×10^10^ (Purple), 1×10^9^ (light green), 1×10^8^ (dark green), 1×10^7^ (light blue), 1×10^6^ (bordeaux), 1×10^5^ (green), 1×10^4^ (yellow), 1×10^3^ (orange) and 75 copies (blue). Samples were amplified in triplicates. Average values are shown for each point

B) Linear amplification range of viral RNA. Ct value vs log10 (Copy number per reaction). Each point is the average of three replicates Top-right: linear equation and correlation coefficient. The slope of the curve is -3.30, indicating an amplification efficiency of 1.01 in the range of 15 copies/µl to 2×10^9^ copies/µl.

**Supplementary Table 1**.

Acknowledgments to the authors that generated SARS-CoV-2 genome sequences publicly available at the GISAID database accessed on June 2^nd^ 2020.

